# Assessing avoidant/restrictive food intake disorder symptoms using the Nine Item ARFID Screen in >9,000 Swedish adults with and without eating disorders

**DOI:** 10.1101/2024.05.27.24307888

**Authors:** Emily K Presseller, Gabriella E Cooper, Laura M Thornton, Andreas Birgegård, Afrouz Abbaspour, Cynthia M Bulik, Emma Forsén Mantilla, Lisa Dinkler

## Abstract

**Objective:** The Nine Item ARFID Screen (NIAS) is a widely used measure assessing symptoms of avoidant/restrictive food intake disorder (ARFID). Previous studies suggest that individuals with eating disorders driven by shape/weight concerns also have elevated scores on the NIAS. To further clarify this issue, we characterized NIAS scores in a large sample of individuals with eating disorders and evaluated overlap in symptoms measured by the NIAS and the Eating Disorder Examination-Questionnaire (EDE-Q) version 6.0.

**Method:** Our sample comprised 9,148 participants from the Eating Disorders Genetics Initiative Sweden (EDGI-SE), who completed surveys including NIAS and EDE-Q. NIAS scores were calculated and compared by eating disorder diagnostic group using descriptive statistics and linear models.

**Results:** Participants with current anorexia nervosa demonstrated the highest mean NIAS scores and had the greatest proportion (57.0%) of individuals scoring above a clinical cutoff on at least one of the NIAS subscales. Individuals with bulimia nervosa, binge-eating disorder, and other specified feeding or eating disorder also demonstrated elevated NIAS scores compared to individuals with no lifetime history of an eating disorder (*p*s < .05). All subscales of the NIAS showed small to moderate correlations with all subscales of the EDE-Q (*r*s = 0.26-0.40).

**Discussion:** Our results substantiate that individuals with eating disorders other than ARFID demonstrate elevated scores on the NIAS, suggesting that this tool is inadequate on its own for differentiating ARFID from shape/weight-motivated eating disorders. Further research is needed to inform clinical interventions addressing the co-occurrence of ARFID-related drivers and shape/weight-related motivation for dietary restriction.

## Introduction

First published in 2018, the Nine Item ARFID Screen (NIAS) was the first self-reported questionnaire specifically developed to assess symptoms of avoidant restrictive food intake disorder (ARFID; Zickgraf & Ellis, 2018). The NIAS is comprised of nine items that are scored on a 6-point Likert scale from *Strongly disagree* (0) to *Strongly agree* (5). It yields a total score and three subscale scores that identify different dimensions of ARFID, including picky eating (“I am a picky eater”), low appetite (“I am not very interested in eating; I seem to have a smaller appetite than other people”), and fear of aversive consequences (“I eat small portions because I am afraid of GI [gastrointestinal] discomfort, choking, or vomiting”). Initial validation of the NIAS indicated high internal consistency; test-retest reliability; factor loading invariance; convergent validity with other self-report measures of picky eating (specifically, the Dietary Variety Questionnaire, Food Fussiness subscale of the Adult Eating Behavior Questionnaire, and Food Neophobia Scale), appetite (Satiety Responsiveness, Emotional Under-eating, and Slowness in Eating subscales of the Adult Eating Behavior Questionnaire), and fear of aversive consequences (Emetophobia Questionnaire and Visceral Sensitivity Index); and discriminant validity with other forms of psychopathology (Zickgraf & Ellis, 2018).

The NIAS has been widely used in a number of settings with diverse clinical and non-clinical populations (Zickgraf et al., 2023) and norms for the general population have been established (Zickgraf & Ellis, 2018). A large number of translations and adaptations of the NIAS have been made (e.g., Billman Miller et al., 2024; Fekih-Romdhane et al., 2023; He et al., 2021; Medina-Tepal et al., 2023; Van Ouytsel et al., 2024; Ziolkowska et al., 2022), including development of a parent-report version (Ziolkowska et al., 2022). The NIAS has also been used to examine the potential comorbidity of ARFID in individuals with gastrointestinal disorders, including gastroparesis, functional dyspepsia, achalasia, celiac disease, eosinophilic esophagitis, and inflammatory bowel disease (Burton Murray et al., 2020; Fink et al., 2022; Kaul et al., 2024; Robelin et al., 2021; Yelencich et al., 2022).

However, several smaller clinical studies have now shown that the NIAS does not seem to differentiate well between ARFID and eating disorders driven by shape/weight concerns, because individuals with other eating disorders also have elevated scores on the NIAS (Billman Miller et al., 2024; Burton Murray et al., 2021). These studies found that the NIAS alone was insufficient to distinguish accurately between individuals with ARFID and individuals with other eating disorders, suggesting that the NIAS may capture both ARFID-related eating disturbances and eating behaviors associated with weight/shape driven eating disorders (Billman Miller et al., 2024; Burton Murray et al., 2021). Accordingly, findings from these studies have yielded recommendations to combine the NIAS with additional eating disorder questionnaires as exclusion measures when assessing ARFID symptoms.

Furthermore, although presence of other eating disorder pathology is theoretically excluded by the ARFID diagnostic criteria defined by the Diagnostic and Statistical Manual of Mental Disorders, Fifth Edition (DSM-5; American Psychiatric Association, 2013), clinical observations indicate that patients with clear ARFID *do* sometimes present with symptoms of other eating disorders (e.g., binge eating, weight and shape concerns), further complicating the issue. Becker et al. (2020) described two cases of ARFID symptomatology that overlapped with traditional eating disorder symptoms, including fear of weight gain, influence of shape and weight on self-evaluation, caloric restriction, fasting, and binge eating. Similarly, Barney et al. (2022) reported a case of ARFID in a 9-year-old girl who also endorsed thin ideal internalization and body image concerns. In this case, the authors argued that ARFID was still the appropriate diagnosis for this individual, given that sensory aversion and low appetite were the primary factors driving engagement in food restriction. In a cohort of patients in day treatment for eating disorders, Nicely et al. (2014) reported that 21% of patients with ARFID also experienced concerns related to their shape and/or weight, including concerns about physical health risk and body dissatisfaction related to *low* weight.

Thus, the aim of the present study was to further assess the usefulness of the NIAS within a large sample (N > 9,000) of people with diverse current and previous eating disorders. We compared ARFID symptoms (i.e., the distribution of NIAS scores) in people with current anorexia nervosa (AN), bulimia nervosa (BN), binge-eating disorder (BED), and other specified feeding or eating disorders (OSFED); people with previous eating disorders; and people without current or previous eating disorders. We also explored overlap in symptoms measured by the NIAS and the Eating Disorder Examination-Questionnaire (EDE-Q) among these individuals. Considering that both AN and ARFID are restrictive eating disorders and therefore have the largest behavioral symptom overlap, we hypothesized that ARFID symptoms would be highest among individuals with current AN.

## Method

### Participants

The study sample included individuals who participated in the Eating Disorders Genetics Initiative Sweden (EDGI-SE). EDGI-SE has ethical approval from the Swedish Ethical Review Authority (dnr 2020-02243). Eligible participants for EDGI-SE were individuals aged 16 years and older, with current or previous experience of eating disorders (cases) or without any such experience (controls). Participants had to have an address in Sweden, have a Swedish Bank ID (electronic personal identifier), and be able to understand Swedish. There were no further exclusion criteria for cases. Controls and sub-threshold controls were excluded if they stated that they had indeed had an eating disorder; that they had been hospitalized for some other serious psychiatric illness (e.g., bipolar disorder, schizophrenia), or that a first-degree relative had had an eating disorder. Recontacted individuals who scored as controls in previous studies but now scored as cases were also excluded.

### Procedure

The recruitment approach was multi-pronged with participants recruited via participation in previous studies, public outreach, the Swedish national quality register for eating disorders (Riksät), and the market research company IPSOS. Participants from previous studies and Riksät were sent an invitation to participate in EDGI via e-mail or post. Public outreach included posts in social media channels; promotion in podcasts, magazine and newspaper articles; information about EDGI in a large production on Swedish national television; advertisement in the member magazine of the largest Swedish eating disorder patient advocacy organization; and, lastly, posters and folders at several eating disorder clinics around the country. Trained interviewers from the company IPSOS primarily recruited controls, and potential participants were contacted by phone. Interviewers screened for eligibility and those who were interested were sent further information and an invitation to participate via e-mail.

Individuals opting to participate in EDGI-SE followed the same procedure for providing consent and completing the surveys. They signed up on the official project website (www.edgi.se) and, after receiving the appropriate information, they consented using BankID (a method for secure identification in Sweden). After consent, participants provided updated contact details and completed online screening. Some individuals (n=458) were excluded based on the screening results, but those who were included as cases, controls, or sub-threshold controls then progressed to the online main survey. The screening survey took about 15 minutes and the main survey about 30 minutes, with some variation depending on group (cases answered a few more questionnaires than controls). After completing the main survey, participants who had not provided saliva in previous studies were sent kits for saliva DNA sampling (OG-500). However, since the genomic data are not included in the present analyses, this part of the procedure is not described further. After completing all steps above, participants were sent a thank you letter, including two cinema gift cards as a token of appreciation.

### Measures

In total, EDGI-SE comprises 14 surveys (1 for screening, 13 in the main survey). The main battery is almost identical to that used at other international EDGI sites and has been described elsewhere (Bulik et al., 2021). Current ARFID symptoms were assessed with the NIAS, which was described in detail in the introduction.

The algorithm for identifying eating disorder status is shown in **Table 1**. The presence or absence of<u>current</u> eating disorders was identified using the diagnostic definitions proposed by Berg et al. (2012) which are based on EDE-Q 6.0 (Fairburn & Beglin, 2008; Welch et al., 2011). EDE-Q 6.0 is a 28-item self-report assessing current cognitive and behavioral eating disorder pathology, summarized in a Global Score and four subscales: Restraint, Eating Concerns, Weight Concerns and Shape Concerns. The behavioral items ask about presence and frequency of common eating disorder behaviors (such as binge eating and compensatory behaviors) with the respondent specifying how often they have occurred over the past 28 days, with 0 indicating not at all. The other items are rated on a 7-point Likert scale, with higher scores indicating greater difficulties.

**Table 1.** Identification of current eating disorder status (operationalization in grey italics)

| Diagnosis | (A) Fear of Weight Gain | (B) Importance of Weight | (C) Importance of Shape | (D) Binge eating | (E) Purging | (F) Current BMI | (G) Current ED Status | (H) Lifetime ED (EDGI-SE group) | Condition |
| --- | --- | --- | --- | --- | --- | --- | --- | --- | --- |
| **Current AN** | Rated $\geq 4$ in the last 28 days: *EDE-Q item 10 $\geq 4$* | Rated $\geq 4$ in the last 28 days: *EDE-Q item 22 $\geq 4$* | Rated $\geq 4$ in the last 28 days: *EDE-Q item 23 $\geq 4$* | ---- | $\geq 4$ purging episodes in the last 28 days: *Sum of EDE-Q items 16, 17, 18 $\geq 4$* | $< 18.55$ | ---- | --- | (A or E) + (B or C) + F |
| **Current BN** | --- | Rated $\geq 4$ in the last 28 days: *EDE-Q item 22 $\geq 4$* | Rated $\geq 4$ in the last 28 days: *EDE-Q item 23 $\geq 4$* | $\geq 4$ binge-eating episodes in the last 28 days: *EDE-Q item 15 $\geq 4$* | $\geq 4$ purging episodes in the last 28 days: *Sum of EDE-Q items 16, 17, 18 $\geq 4$* | $\geq 18.55$ | ---- | --- | (B or C) + D + E + F |
| **Current BED** | --- | --- | --- | $\geq 4$ binge-eating episodes in the last 28 days: *EDE-Q item 15 $\geq 4$* | $\leq 1$ episode of purging in the last 28 days: *Sum of EDE-Q items 16, 17, 18 $\leq 1$* | $\geq 18.55$ | ---- | --- | D + E + F |
| **Current OSFED** | --- | --- | --- | --- | --- | --- | No current AN, BN, or BED (as defined above) + does not meet criteria for Never ED (as defined below) | --- | G |
| **Previous ED** | Rated $\leq 2$ in the last 28 days: *EDE-Q item 10 $\leq 2$* | Rated $\leq 2$ in the last 28 days: *EDE-Q item 22 $\leq 2$* | Rated $\leq 2$ in the last 28 days: *EDE-Q item 23 $\leq 2$* | 0 binge-eating episodes in the last 28 days: *EDE-Q item 15 = 0* | 0 episodes of purging in the last 28 days: *Sum of EDE-Q items 16, 17, 18 = 0* | ---- | Does not meet criteria for any current ED diagnosis (as defined in Current AN, Current BN, Current BED, and Current OSFED rows) | Lifetime ED Case in EDGI-SE | A + B + C + D + E + G + H |
| **Never ED** | Rated $\leq 2$ in the last 28 days: *EDE-Q item 10 $\leq 2$* | Rated $\leq 2$ in the last 28 days: *EDE-Q item 22 $\leq 2$* | Rated $\leq 2$ in the last 28 days: *EDE-Q item 23 $\leq 2$* | 0 binge-eating episodes in the last 28 days: *EDE-Q item 15 = 0* | 0 episodes of purging in the last 28 days: *Sum of EDE-Q items 16, 17, 18 = 0* | ---- | Does not meet criteria for any current ED diagnosis (as defined in Current AN, Current BN, Current BED, and Current OSFED rows) | No lifetime ED Control in EDGI-SE | A + B + C + D + E + G + H |

AN: anorexia nervosa, BN: bulimia nervosa, BED: binge-eating disorder, OSFED: other specified feeding or eating disorder, BMI: body mass index, ED: eating disorder, EDGI-SE: Eating Disorders Genetics Initiative-Sweden, EDE-Q: Eating Disorder Examination-Questionnaire
EDE-Q item 10: "Have you had a definite fear that you might gain weight?"; response options: no days (0), 1-5 days (1), 6-12 days (2), 13-15 days (3), 16-22 days (4), 23-27 days (5), every day (6)
EDE-Q item 15: Over the past 28 days, on how many DAYS have such episodes of overeating occurred (i.e. you have eaten an unusually large amount of food and have had a sense of loss of control at the time)?"; response=frequency
EDE-Q item 16: "Over the past 28 days, how many times have you made yourself sick (vomit) as a means of controlling your shape or weight?"; response=frequency
EDE-Q item 17: "Over the past 28 days, how many times have you taken laxatives as a means of controlling your shape or weight?"; response=frequency
EDE-Q item 18: "Over the past 28 days, how many times have you exercised in a "driven" or "compulsive" way as a means of controlling your weight, shape or amount of fat, or to burn off calories?"; response=frequency
EDE-Q item 22: "Over the past 28 days, has your weight influenced how you think about (judge) yourself as a person?"; response options: no days (0), 1-5 days (1), 6-12 days (2), 13-15 days (3), 16-22 days (4), 23-27 days (5), every day (6)
EDE-Q item 23: "Over the past 28 days, has your shape influenced how you think about (judge) yourself as a person?"; response options: no days (0), 1-5 days (1), 6-12 days (2), 13-15 days (3), 16-22 days (4), 23-27 days (5), every day (6)

As EDE-Q only collects information about current eating disorder symptoms (past 28 days), more information was needed to differentiate the *previous* eating disorder and *never* eating disorder groups. Within the EDGI-SE parent study, *lifetime* EDGI participant categories were determined using ED100K, in accordance with previously published EDGI methods (Thornton et al., 2018): (1) individuals with a history of or a current eating disorder (cases), (2) individuals with a history of or a current *sub-threshold* eating disorder (sub-threshold cases), and (3) individuals with *no history* of eating disorder (controls; **Table 2**). In the current study, lifetime EDGI participant categories were used to identify individuals with *previous* (but not current) eating disorder (EDGI case but no current eating disorder according to Berg et al.’s EDE-Q diagnostic algorithm) and individuals in the *never* eating disorder group (EDGI control and no current eating disorder according to Berg et al.’s EDE-Q diagnostic algorithm; see **Table 1**).

**Table 2.** Sample (N = 9,148) demographic and clinical characteristics.

| Characteristic | *N* = 9,148 |  |
| --- | --- | --- |
|  | *Mean (SD)* | *Median (Min-Max)* |
| **Age** | 32.1 (11.0) | 30 (15-79) |
| **BMI** | 23.5 (5.5) | 22.2 (6.3-74.2) |
|  | *n* | *%* |
| **Sex** |  |  |
| Male | 267 | 2.9% |
| Female | 8,880 | 97.1% |
| **Gender** |  |  |
| Male | 286 | 3.1% |
| Female | 8,748 | 95.6% |
| Non-binary | 102 | 1.1% |
| Other | 12 | 0.1% |
| **EDGI-SE category** |  |  |
| Case | 8,030 | 87.8% |
| Control | 880 | 9.6% |
| Sub-threshold case | 238 | 2.6% |
| **Lifetime AN** | 5,601 | 61.2% |
| **Lifetime BN** | 2,208 | 24.1% |
| **Lifetime BED** | 833 | 9.1% |
| **Lifetime Atypical AN** | 1,287 | 14.1% |
| **Lifetime Atypical BN** | 720 | 7.9% |
| **Lifetime Atypical BED** | 369 | 4.0% |
| **Current ED status** |  |  |
| Current AN | 484 | 5.3% |
| Current BN | 699 | 7.6% |
| Current BED | 643 | 7.0% |
| Current OSFED | 4,523 | 49.4% |
| Previous ED | 2,004 | 21.9% |
| Never ED | 795 | 8.7% |

SD: standard deviation, Min: minimum, Max: maximum, BMI: body mass index, EDGI-SE: Eating Disorders Genetics Initiative Sweden, AN: anorexia nervosa, BN: bulimia nervosa, BED: binge-eating disorder, ED: eating disorder, OSFED: other specified eating disorder. When data was collected for only a subset of the total sample, *N* is specified in parenthetical next to variable label. Lifetime eating disorder diagnoses reflect the frequencies of each diagnosis at any point in the participants' lives and accordingly, participants may belong to multiple lifetime eating disorder groups.

### Statistical Analysis

All statistical analyses were conducted in R version 4.3.1. First, we examined the distribution of continuous NIAS total score, subscale scores, and individual item scores by eating disorder groups using linear regression. Pairwise estimated marginal means with a family-wise error rate adjustment (Bonferroni correction) were used to compare specific eating disorder groups. Cohen’s d effect sizes were computed for pairwise comparisons and only medium and large effects were interpreted as clinically meaningful. Second, we compared the proportion scoring above the empirically derived NIAS cutoff scores (NIAS Picky eating >= 10, NIAS Appetite >= 9, and NIAS Fear >= 10, respectively; Burton Murray et al., 2021) between eating disorder groups using Pearson’s chi-squared test.

## Results

Our sample comprised N = 9,148 participants (2.9% male) with EDE-Q data necessary for eating disorder group assignment and at least one complete NIAS subscale score. **Table 2** displays demographic characteristics and the prevalence of current eating disorder diagnoses according to the EDE-Q. **Table 2** also includes information about distribution of eating disorder diagnoses in our sample across their lifetimes according to the ED100K. More than half of the sample (61.2%) had AN at some point in their lifetime. The majority (n=6,349; 69.4%) had a *current* eating disorder; the most common current eating disorder was OSFED (49.4%).

**Table 3** depicts the proportion of participants scoring above NIAS clinical cutoff scores by eating disorder diagnosis. More than 50% of participants with current AN scored above at least one of the NIAS clinical cutoff scores, followed by 30% of participants with current BN, 23% of participants with current OSFED, and 17% of participants with current BED. Of the NIAS subscales, participants with current AN or current OSFED were most likely to score above the clinical cutoff on the NIAS Appetite subscale, whereas participants with current BN or current BED were most likely to score above the clinical cutoff on the NIAS Picky Eating subscale.

**Table 3.** Distribution of dichotomized NIAS scores by current eating disorder status, n (%)

| Characteristic | Overall<br>N = 9,148 | Current AN<br>N = 484 | Current BN<br>N = 699 | Current BED<br>N = 643 | Current OSFED<br>N = 4,523 | Previous ED<br>N = 2,004 | Never ED<br>N = 795 | p-value<sup>1</sup> |
| --- | --- | --- | --- | --- | --- | --- | --- | --- |
| Any NIAS subscale score above cutoff | 1,858 (20.3%) | 276 (57.0%) | 212 (30.3%) | 107 (16.6%) | 1,060 (23.4%) | 195 (9.7%) | 8 (1.0%) | <0.001 |
| NIAS Picky eating score $\geq 10$ | 971 (10.6%) | 130 (26.9%) | 131 (18.7%) | 73 (11.4%) | 558 (12.3%) | 73 (3.6%) | 6 (0.8%) | <0.001 |
| NIAS Appetite score $\geq 9$ | 1,128 (12.3%) | 215 (44.4%) | 82 (11.7%) | 34 (5.3%) | 666 (14.7%) | 129 (6.4%) | 2 (0.3%) | <0.001 |
| NIAS Fear score $\geq 10$ | 421 (4.6%) | 72 (14.9%) | 59 (8.4%) | 22 (3.4%) | 232 (5.1%) | 36 (1.8%) | 0 (0.0%) | <0.001 |

<sup>1</sup>Pearson's chi-squared test
NIAS: Nine Item ARFID Screen, AN: anorexia nervosa, BN: bulimia nervosa, BED: binge-eating disorder, OSFED: other specified feeding or eating disorder, ED: eating disorder

**Table 4** and **Figure 1** depict the distribution of NIAS total, subscale, and item-level scores by eating disorder group. The current AN group demonstrated the highest average NIAS total score, followed by BN, OSFED, and BED and there was a significant difference between eating disorder groups on NIAS total and subscale scores. The AN group demonstrated significantly higher scores than the BN, BED, and OSFED groups on the NIAS total and Appetite subscale scores with large effect sizes (see Table S1). The AN group demonstrated significantly higher scores on the Picky Eating and Fear subscales than the BED and OSFED groups with medium to large effect sizes. Although the AN group had significantly higher scores than the BN group on the Picky Eating and Fear subscales, the effect sizes for these pairwise comparisons were small. The BN group demonstrated significantly higher scores than the BED group on the Fear subscale with medium effect size. All other pairwise comparisons between threshold eating disorder groups on NIAS total and subscale scores were of small effect sizes. Mean scores on individual items of the NIAS demonstrated a similar pattern across eating disorder groups as the NIAS total and subscale scores (see **Tables 4 and S1**). Within the Picky Eating subscale, “I am a picky eater” was the item with the highest mean score across the overall sample. “I have to push myself to eat regular meals throughout the day, or to eat a large enough amount of food at meals” was the highest rated item on the Appetite subscale. On the Fear subscale, “I restrict myself to certain foods because I am afraid that other foods will cause GI (gastrointestinal) discomfort, choking, or vomiting” had the highest mean rating of all subscale items.

**Figure 1.**
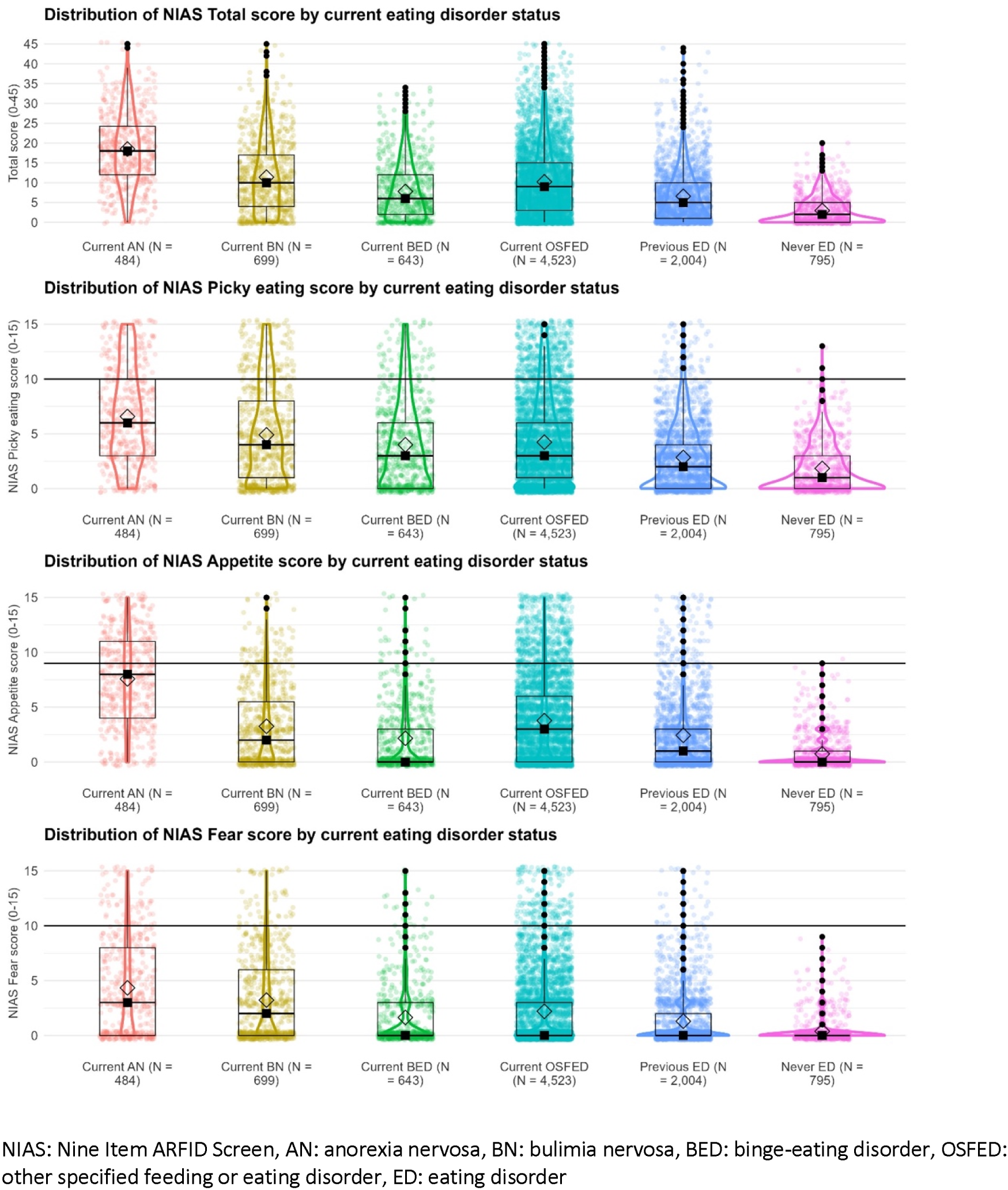
Distribution of NIAS scores by current eating disorder status

**Table 4.**
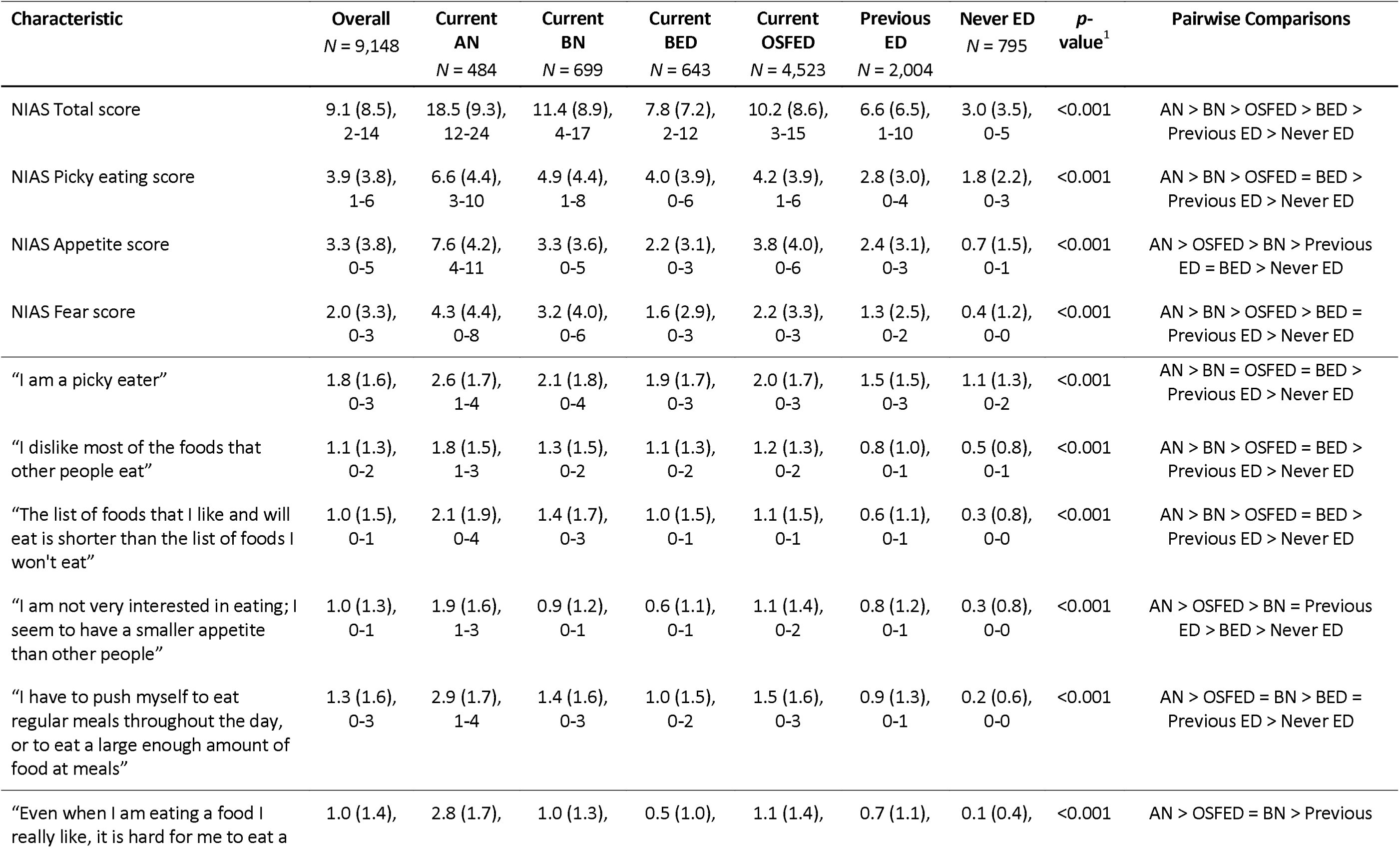
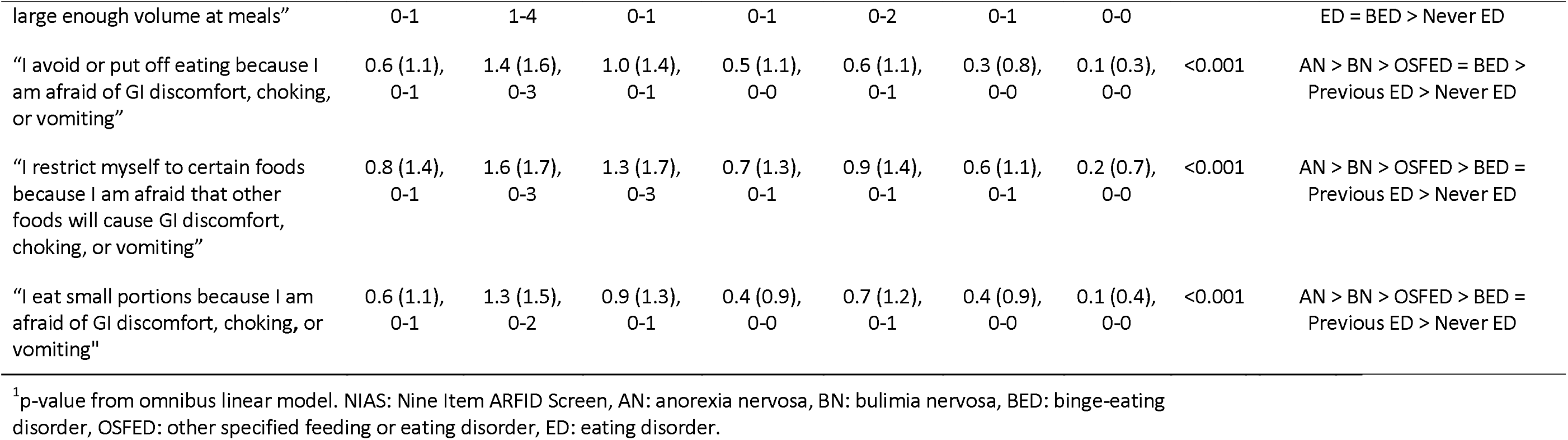
Distribution of continuous NIAS scores and single NIAS items by current eating disorder status, Mean (SD), IQR.

**Table 5** depicts the correlations between EDE-Q total and subscale scores and NIAS total and subscale scores across the complete sample. All correlations were of medium effect size (approximately *r* = 0.30 or greater).

**Table 5.**
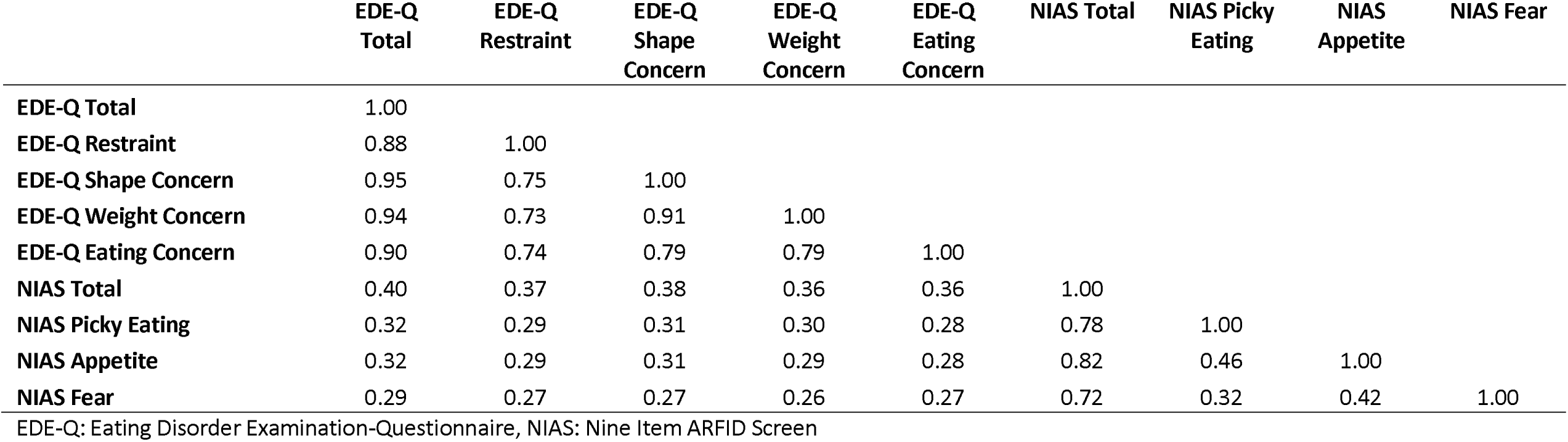
Correlations of EDE-Q scores with NIAS scores (all significant at p<.001)

|  | **EDE-Q**<br>**Total** | **EDE-Q**<br>**Restraint** | **EDE-Q**<br>**Shape**<br>**Concern** | **EDE-Q**<br>**Weight**<br>**Concern** | **EDE-Q**<br>**Eating**<br>**Concern** | **NIAS Total** | **NIAS Picky**<br>**Eating** | **NIAS**<br>**Appetite** | **NIAS Fear** |
| --- | --- | --- | --- | --- | --- | --- | --- | --- | --- |
| **EDE-Q Total** | 1.00 |  |  |  |  |  |  |  |  |
| **EDE-Q Restraint** | 0.88 | 1.00 |  |  |  |  |  |  |  |
| **EDE-Q Shape Concern** | 0.95 | 0.75 | 1.00 |  |  |  |  |  |  |
| **EDE-Q Weight Concern** | 0.94 | 0.73 | 0.91 | 1.00 |  |  |  |  |  |
| **EDE-Q Eating Concern** | 0.90 | 0.74 | 0.79 | 0.79 | 1.00 |  |  |  |  |
| **NIAS Total** | 0.40 | 0.37 | 0.38 | 0.36 | 0.36 | 1.00 |  |  |  |
| **NIAS Picky Eating** | 0.32 | 0.29 | 0.31 | 0.30 | 0.28 | 0.78 | 1.00 |  |  |
| **NIAS Appetite** | 0.32 | 0.29 | 0.31 | 0.29 | 0.28 | 0.82 | 0.46 | 1.00 |  |
| **NIAS Fear** | 0.29 | 0.27 | 0.27 | 0.26 | 0.27 | 0.72 | 0.32 | 0.42 | 1.00 |

EDE-Q: Eating Disorder Examination-Questionnaire, NIAS: Nine Item ARFID Screen

## Discussion

The present study characterized NIAS scores among a large sample of Swedish adults with current and previous eating disorders. Results suggest that people with current eating disorders other than ARFID have elevated scores on the NIAS, generally exceeding scores among individuals with previous eating disorders and control individuals with no current or previous eating disorders. Individuals with current AN demonstrated significantly and clinically meaningfully higher scores than individuals with current BED and OSFED across all subscales of the NIAS but differed from individuals with BN in a clinically meaningful way only on the NIAS total score and Appetite subscale. Further, a substantial proportion of individuals with current eating disorders scored above clinical cutoff scores, especially on the Picky Eating and Appetite subscales. In particular, individuals with current AN were most likely to score above clinical cutoffs on the NIAS, potentially highlighting the multiple reasons for which individuals with current AN may endorse dietary restriction beyond shape and weight concerns. Care should be taken to comprehensively assess primary and additional motivations for dietary restriction among individuals presenting with low weight AN, especially given that the ego-syntonic nature of AN may discourage individuals with AN from endorsing shape and weight concerns (Guarda, 2008).

The results of this study replicate previous findings suggesting that the NIAS alone is insufficient for differentiating ARFID from other eating disorder presentations (Billman Miller et al., 2024; Burton Murray et al., 2021). Our findings quantify this problem and contribute new insight into the relative elevation of individual NIAS subscale scores among individuals with different types of eating disorders, highlighting that individuals with AN are more likely than individuals with other current eating disorders to have elevated scores on the NIAS, especially the Appetite subscale. The most ARFID-specific subscale of the NIAS might be the Fear of aversive consequences subscale, as only 4.6% of our sample scored above the clinical cutoff on this subscale, compared to 12.3% on Appetite and 10.6% on Picky Eating. However, this subscale only captures the portion of individuals with ARFID whose presentation is related to fear of aversive consequences. Further, research suggests that fear of aversive consequences is the least common ARFID presentation, making it even more problematic that the Fear subscale of the NIAS seems to be most ARFID-specific, as it likely only captures a small fraction of individuals with ARFID (Eddy et al., 2015; Fisher et al., 2014; Kurz et al., 2015). On the other hand, GI pain and discomfort are also common in eating disorders other than ARFID and concern about GI symptoms may lead to restriction of food intake, particularly in anorexia nervosa (Zucker & Bulik, 2020). Therefore, fear of choking and fear of vomiting that might be the most specific to ARFID. However, different types of fear of aversive consequences are not distinguishable in the NIAS items. To reliably identify people with ARFID, another measure of eating pathology such as the EDE-Q is essential to rule out the presence of other eating pathology and avoid identifying individuals with other eating disorders as having ARFID. Interestingly, within our sample, all EDE-Q subscale scores demonstrated moderate correlations with NIAS subscale scores.

Our findings also support a growing body of literature suggesting that there may be more symptom overlap between ARFID and other eating disorders than described within the Diagnostic and Statistical Manual for Mental Disorders, Fifth Edition (DSM-5; American Psychiatric Association, 2013). Among individuals with restrictive eating, reported motivations for insufficient dietary intake may include low appetite and dislike of specific foods alongside shape- and weight-related motivations (Kambanis, Mulkens, et al., 2024). This may complicate appropriate diagnosis of these individuals, yielding downstream effects on case conceptualization and treatment planning. For instance, if an individual with AN is inappropriately diagnosed with ARFID, body image concerns may be de-emphasized during treatment, increasing this individual’s potential for post-treatment relapse of dietary restriction. Conversely, given the “normative discontent” that most individuals experience regarding body image, individuals with ARFID (i.e., dietary restriction primarily driven by factors other than shape or weight concerns) may be inappropriately diagnosed with a traditional eating disorder and treatment may place too much emphasis on addressing shape and weight concerns, yielding poorer treatment response (Tantleff-Dunn et al., 2011) and potentially causing an iatrogenic weight or shape driven eating disorder. Further complicating the clinical picture for some patients, emerging evidence suggests that shape and weight concerns and other eating pathology may emerge following onset of ARFID, yielding diagnostic crossover among some patients (Kambanis, Mancuso, et al., 2024). These data may indicate a need to update extant treatment approaches to address both shape and weight *and* ARFID-related motivations for dietary restriction among transdiagnostic feeding/eating disorder presentations. Additional studies with larger samples and longer follow-ups are required to confidently determine the extent to which this diagnostic migration occurs.

The present study had several strengths and limitations. Our study advances understanding of the NIAS by replicating previous findings in a largely non-clinical sample of individuals with different types of current and previous eating disorders, and controls with no eating disorder history. Our study was limited by not recruiting individuals with ARFID or including more extensive ARFID diagnostic instruments in the assessment. We were therefore unable to identify what proportion of our sample may have met or may currently meet DSM-5 criteria for ARFID, nor could we compare NIAS scores among individuals in the EDGI-SE sample with ARFID to participants with other current eating disorders. We also only used self-report measures of ARFID symptoms (the NIAS) and other eating disorder symptoms (the EDE-Q, ED100K), which are likely limited by participant insight, demand characteristics, and recall biases. Large studies such as EDGI-SE must make difficult choices between sample size, which is essential for genetic studies, and method of assessment, with diagnostic interviews on such large samples being untenable.

Future research should continue to explicate the symptom overlap and diagnostic crossover between ARFID and other eating disorder diagnoses and subsequent diagnostic modifications should consider these findings when determining whether to allow for co-occurrence of ARFID and other eating disorders. If crossover from ARFID to other eating disorders (or vice versa) does indeed occur, its course and predictors should be identified. Adaptations to extant treatment approaches for ARFID and other eating disorders may be necessary to comprehensively treat individuals whose drivers of eating pathology may cross existing diagnostic borders.

## Data Availability

Data are available on request from the authors.

## Public Significance Statement

Differentiating symptoms of avoidant/restrictive food intake disorder (ARFID) from symptoms of other eating disorders continues to be a challenge for the field. This study demonstrated that a substantial proportion of individuals with current eating disorders (e.g., anorexia nervosa, bulimia nervosa, binge-eating disorder) score above clinical cutoff for ARFID on the Nine Item ARFID Screen, suggesting that this tool is inadequate on its own for differentiating ARFID from shape/weight-motivated eating disorders.

## Funding statement or other acknowledgements of support

We gratefully acknowledge the contribution of the participants Eating Disorder Genetics Initiative-Sweden (EDGI-SE). EDGI-SE was funded by the Swedish Research Council (Vetenskapsrådet, 538-2013-8864, PI: Bulik). Cynthia M. Bulik was supported by NIMH (R56MH129437; R01MH120170; R01MH124871; R01MH119084; R01MH118278); Swedish Research Council (538-2013-8864); Lundbeck Foundation (R276-2018-4581). Lisa Dinkler was supported by Swedish Society for Medical Research (SSMF, PG-22-0478), Swedish Mental Health Foundation (Fonden för Psykisk Hälsa 2022 & 2023); and Fredrik and Ingrid Thurings Foundation (Thurings stiftelse 2021-00660). Emily K. Presseller was supported by NIMH (F31MH131262). The funders had no role in the design/conduct of the study, data management/analysis, or manuscript preparation.

## Data, materials and code availability statement

Data are available on request from the authors.

## Authors’ contributions

Conceptualization: Emily K Presseller, Gabriella E Cooper, Cynthia M. Bulik, Lisa Dinkler

Data curation: Emily K Presseller, Gabriella E Cooper, Andreas Birgegård, Emma Forsén Mantilla, Lisa Dinkler

Formal analysis: Emily K Presseller, Gabriella E Cooper, Lisa Dinkler

Funding acquisition: Cynthia M. Bulik. Lisa Dinkler

Investigation: Laura M Thornton, Cynthia M. Bulik, Emma Forsén Mantilla

Methodology: Emily K Presseller, Gabriella E Cooper, Cynthia M. Bulik, Lisa Dinkler

Project administration: Lisa Dinkler

Resources: Laura M Thornton, Afrouz Abbaspour, Cynthia M. Bulik, Emma Forsén Mantilla

Software: Emily K Presseller, Gabriella E Cooper, Lisa Dinkler

Supervision: Lisa Dinkler

Validation: Emily K Presseller, Lisa Dinkler

Visualization: Emily K Presseller, Lisa Dinkler

Writing – original draft: Emily K Presseller, Gabriella E Cooper, Liv Hog, Lisa Dinkler

Writing – review & editing: All authors

