## Supplemental file for "Assessing avoidant/restrictive food intake disorder symptoms using the Nine Item ARFID Screen in >9,000 Swedish adults with and without eating disorders"

**Table S1.** Pairwise comparison and effect sizes (Cohen’s *d*) for NIAS subscales and items by current ED status

|  | **Pairwise Cohen’s d** | | | | | | | | | | | | | | | | | | | | | | | | | | | | |
| --- | --- | --- | --- | --- | --- | --- | --- | --- | --- | --- | --- | --- | --- | --- | --- | --- | --- | --- | --- | --- | --- | --- | --- | --- | --- | --- | --- | --- | --- |
|  | AN-BN | | AN-BED | | AN-OSFED | | AN-Previous ED | | AN-Never ED | | BN-BED | | BN-OSFED | | BN-Previous ED | | BN-Never ED | | BED-OSFED | | BED-Previous ED | | BED-Never ED | | OSFED Previous ED | | OSFED-Never ED | | Previous ED-Never ED |
| NIAS Total score | **0.91** | | **1.36** | | **1.06** | | **1.52** | | **1.98** | | 0.46 | | 0.15 | | **0.61** | | **1.08** | | 0.31 | | 0.16 | | **0.62** | | 0.46 | | **0.93** | | 0.46 |
| NIAS Picky eating score | 0.46 | | **0.70** | | **0.64** | | **1.02** | | **1.29** | | 0.25 | | 0.18 | | **0.56** | | **0.83** | | -0.06 | | 0.31 | | **0.59** | | 0.38 | | **0.65** | | 0.27 |
| NIAS Appetite score | **1.21** | | **1.52** | | **1.06** | | **1.44** | | **1.92** | | 0.31 | | -0.15 | | 0.23 | | **0.70** | | -0.45 | | 0.08 | | 0.40 | | 0.38 | | **0.85** | | 0.47 |
| NIAS Fear score | 0.36 | | **0.86** | | **0.68** | | **0.97** | | **1.26** | | **0.51** | | 0.33 | | **0.61** | | **0.91** | | 0.18 | | 0.11 | | 0.40 | | 0.29 | | **0.58** | | 0.29 |
| “I am a picky eater” | 0.32 | | 0.46 | | 0.40 | | **0.71** | | **0.95** | | 0.15 | | 0.09 | | 0.39 | | **0.63** | | -0.06 | | 0.24 | | 0.49 | | 0.30 | | **0.55** | | 0.24 |
| “I dislike most of the foods that other people eat” | 0.41 | | **0.60** | | **0.54** | | **0.87** | | **1.13** | | 0.20 | | 0.14 | | 0.47 | | **0.73** | | -0.06 | | 0.27 | | **0.53** | | 0.33 | | **0.59** | | 0.26 |
| “The list of foods that I like and will eat is shorter than the list of foods I won't eat” | 0.47 | | **0.77** | | **0.73** | | **1.08** | | **1.28** | | 0.30 | | 0.26 | | **0.61** | | **0.81** | | -0.04 | | 0.30 | | **0.51** | | 0.35 | | **0.55** | | 0.21 |
| “I am not very interested in eating; I seem to have a smaller appetite than other people” | **0.83** | | **1.00** | | **0.61** | | **0.85** | | **1.22** | | 0.17 | | -0.22 | | 0.02 | | 0.39 | | -0.39 | | -0.15 | | 0.22 | | 0.24 | | **0.61** | | 0.37 |
| “I have to push myself to eat regular meals throughout the day, or to eat a large enough amount of food at meals” | **0.98** | | **1.25** | | **0.92** | | **1.31** | | **1.77** | | 0.28 | | -0.06 | | 0.33 | | **0.79** | | -0.33 | | 0.05 | | **0.51** | | 0.39 | | **0.85** | | 0.46 |
| “Even when I am eating a food I really like, it is hard for me to eat a large enough volume at meals” | **1.40** | | **1.75** | | **1.28** | | **1.64** | | **2.05** | | 0.36 | | -0.12 | | 0.24 | | **0.65** | | -0.47 | | 0.12 | | 0.29 | | 0.36 | | **0.77** | | 0.41 |
| “I avoid or put off eating because I am afraid of GI discomfort, choking, or vomiting” | 0.35 | | **0.81** | | **0.70** | | **0.96** | | **1.21** | | 0.45 | | 0.34 | | **0.61** | | **0.86** | | -0.11 | | 0.16 | | 0.41 | | 0.27 | | **0.52** | | 0.25 |
| “I restrict myself to certain foods because I am afraid that other foods will cause GI discomfort, choking, or vomiting” | 0.23 | **0.68** | | **0.53** | | **0.78** | | **1.05** | | 0.45 | | 0.30 | | **0.55** | | **0.83** | | -0.15 | | 0.10 | | 0.38 | | 0.25 | | **0.53** | | 0.27 | |
| “I eat small portions because I am afraid of GI discomfort, choking**,** or vomiting” | **1.04** | **2.51** | | **1.99** | | **2.83** | | **3.68** | | **1.47** | | **0.95** | | **1.79** | | **2.64** | | **-0.53** | | 0.32 | | **1.17** | | **0.84** | | **1.70** | | **0.85** | |

NIAS: Nine Item ARFID Screen, AN: anorexia nervosa, BN: bulimia nervosa, BED: binge-eating disorder, OSFED: other specified feeding or eating disorder, ED: eating disorder. Medium to large effect sizes are bolded to facilitate interpretation.
